# The use of hippocampal grading as a biomarker for early MCI

**DOI:** 10.1101/2022.02.15.22271017

**Authors:** Cassandra Morrison, Mahsa Dadar, Neda Shafiee, D. Louis Collins, Alzheimer’s Disease Neuroimaging Initiative

**Author notes:** Corresponding author: Cassandra Morrison, Montreal Neurological Institute, 3801 University Street, Montreal QC, H3A 2B4. Data used in preparation of this article were obtained from the Alzheimer’s Disease Neuroimaging Initiative (ADNI) database (adni.loni.usc.edu). As such, the investigators within the ADNI contributed to the design and implementation of ADNI and/or provided data but did not participate in analysis or writing of this report. A complete listing of ADNI investigators can be found at: http://adni.loni.usc.edu/wp-content/uploads/how_to_apply/ADNI_Acknowledgement_List.pdf.

## Abstract

**INTRODUCTION:** Changes in the hippocampus are associated with both increased age and cognitive decline due to mild cognitive impairment (MCI) and Alzheimer’s disease (AD). Most studies have examined the association between hippocampal changes and episodic memory, with many reporting a relationship between hippocampal measurements and global cognition. However, these studies often find associations only in the later stages of cognitive decline. The goal of this study was to examine if hippocampal grading is associated with global cognition in cognitively normal controls (NC), early MCI (eMCI), late (lMCI), and AD, and whether such associations differ across diagnostic cohorts.

**METHODS:** Data from 1620 Alzheimer’s Disease Neuroimaging Initiative older adults were examined in this study (495 NC, 262 eMCI, 545 lMCI, and 318 AD). Participants were included if they completed baseline MRI scans and the Alzheimer’s disease Assessment Scale (ADAS-13) and Clinical Dementia Rating – Sum of Boxes (CDR-SB) cognitive tests. Linear regressions examined the influence of hippocampal grading on cognitive scores.

**RESULTS:** Lower global cognition (i.e., increased ADAS-13 scores) was associated with hippocampal grading scores in all cohorts, including normal controls. Lower global cognition (i.e., increased CDR-SB scores) was associated with hippocampal grading scores in lMCI and AD, but not in eMCI or NC groups.

**DISCUSSION:** These findings suggest that hippocampal grading is associated with changes in global cognition in NC, eMCI, lMCI, and AD depending on the cognitive test. Thus, hippocampal grading may be a useful measure that is sensitive to progressive changes early in the disease course.

## 1. Introduction

Increased age is associated with cognitive decline, ranging from healthy aging to mild cognitive impairment (MCI) and, in some cases, Alzheimer’s disease (AD). MCI is characterized by declines in cognitive functioning that are not severe enough to impair daily activites^1^. AD is characterized by progressive declines in cognitive functioning that are severe enough to impair daily activities^2^. In both aging^3^ and AD^4^, neurodegeneration is an important factor that contributes to these observable cognitive deficits.

Hippocampal volume is often used as a biomarker of neurodegeneration because it is affected early in the disease course^5^, with declines in hippocampal volume occurring before observable abnormal amyloid PET^6^. It is well-established that this atrophy occurs decades before the clinical symptom presentation (i.e., measurable cognitive deficits)^7^. For that reason, examining the hippocampus and its relationship to cognitive change in healthy older adults in addition to people with MCI and AD is essential for a detailed understanding of the progressive nature of the disease.

Declines in hippocampal volume are suggested to account for episodic memory declines in aging and AD^8^. Strong associations between decreased hippocampal volume and poor episodic memory functioning support this conclusion^9–11^. In addition to episodic memory, studies have also found a relationship between the hippocampus and global cognitive functioning^12,13^. The correlation between hippocampal volume and global cognition has, however, remained quite moderate, R=0.41^12^. When separating groups based on diagnostic status, global cognition has been correlated with decreased hippocampal volume in only AD (not MCI and NC^14^), and in both AD and MCI (but not NC^15^). Most of these studies used the mini-mental status examination (MMSE) as a measure of global cognition^12,14,15^, which may limit the sensitivity of these associations.

Different measures of the hippocampal neurodegeneration could improve the observed relationship between the hippocampus and general cognitive functioning. Hippocampal grading, measured by the Scoring by Nonlocal Image Patch Estimator (SNIPE), has been shown to surpass hippocampal volume in predictive power^16,17^. These researchers observed that SNIPE-based grading biomarkers are more relevant for cognitive decline prediction and conversion from MCI to AD than hippocampal volume measures^16^. SNIPE has a classification accuracy of 93% at distinguishing people with AD from cognitive normal (CN) individuals in the ADNI cohort^18^. SNIPE could also predict in cognitively normal older adults, who would progress to AD dementia within a 12y follow-up period with 72.5% accuracy^17^. Coupe et al. (2012b) also observed that in normal controls and people with AD, HC grading had a stronger correlation with global cognition, as measured by the MMSE, (R=0.75) than hippocampal volume (R=0.58). Associations between SNIPE hippocampal grading and cognitive declines in normal aging and early MCI have yet to be determined.

Given that the MMSE has limited value in differentiating MCI from NC^19^, other neuropsychological measures that are more sensitive to cognitive changes in early MCI may offer stronger associations with hippocampal volume declines. Furthermore, because hippocampal grading, as measured by SNIPE, has better predictive power than hippocampal volume, we predict that a strong association between SNIPE measures and sensitive measures of cognitive decline will be observed. The current paper was designed to determine whether measures of general cognitive functioning (i.e., the Alzheimer’s Disease Assessment Scale–13, ADAS-13; Clinical Dementia Rating–sum of boxes, CDR-SB) would have a strong association with hippocampal grading in normal controls, people with MCI, and people with AD. While Coupe et al. determined that SNIPE could classify between NC and people with AD and predict which NC may progress to AD, it remains unknown if SNIPE grading is associated with progressive changes in general cognitive functioning at different stages of decline^16–18^. The goal of this paper was to characterize the relationship between SNIPE and cognition as measured by ADAS-13 or CDR-SB early in the disease. Although both ADAS-13 and CDR-SB are sensitive to cognitive changes in MCI and AD, the ADAS-13 has a larger range of scores (0-84) than the CDR-SB (0-18). We expect to see strong associations between the ADAS-13 in NC, MCI, and AD because of the large range in ADAS-13. In contrast, for the CDR-SB we expect to see strong associations only in the later stages of cognitive decline because NCs have a small range of scores (0-0.5). These findings would not only have implications for future development and improvement of methods to measure neurodegeneration, but also for the early detection and characterization of incipient AD.

## 2. Methods

### 2.1 Alzheimer’s Disease Neuroimaging Initiative

Data used in the preparation of this article were obtained from the Alzheimer’s Disease Neuroimaging Initiative (ADNI) database (adni.loni.usc.edu). The ADNI was launched in 2003 as a public-private partnership, led by Principal Investigator Michael W. Weiner, MD. The primary goal of ADNI has been to test whether serial magnetic resonance imaging (MRI), positron emission tomography (PET), other biological markers, and clinical and neuropsychological assessment can be combined to measure the progression of mild cognitive impairment (MCI) and early Alzheimer’s disease (AD). Participants were selected from ADNI-1, ADNI-2, and the ADNI-GO cohorts. The study received ethical approval from the review boards of all participating institutions. Written informed consent was obtained from participants or their study partner.

### 2.2 Participants

Full participant inclusion/exclusion is available online at www.adni-info.org. Briefly, all participants were between 55 and 90 years old at the time of recruitment and no evidence of depression as measured by the Geriatric Depression Scale. Healthy control participants had no evidence of memory decline on the Wechsler Memory Scale and no evidence of cognitive decline on either the Mini Mental Status Examination (MMSE) or Clinical Dementia Rating (CDR). Early MCI and lMCI had to score between 24 and 30 on the MMSE, 0.5 on the CDR, and had abnormal scores on the Wechsler Memory Scale. Early MCI was differentiated from lMCI by degree of memory impairment; eMCI participants were characterized as memory impairment that is intermediate between normal controls and lMCI. AD participants had to show abnormal memory function on the Wechsler Memory Scale, an MMSE score between 20 and 26, a CDR of 0.5 or 1.0 and probable AD according to the NINCDS/ADRDA criteria.

A total of 1634 participants from three ADNI cohorts had MRI baseline scans and were thus included (ADNI-1, 787 participants; ADNI-2, 759 participants; ADNI-GO 88 participants). Inclusion criteria also included availability of baseline CDR-SB and ADAS-13 scores. Fourteen participants were excluded for not having baseline ADAS-13 scores. A total of 1620 participants were included for our study. Of these 1620 participants, 495 were cognitively normal older adults (CN), 262 were early MCI (eMCI), 545 were late MCI (lMCI), and 318 had an AD diagnosis. Table 1 summarizes demographic information for all participants.

**Table 1:**
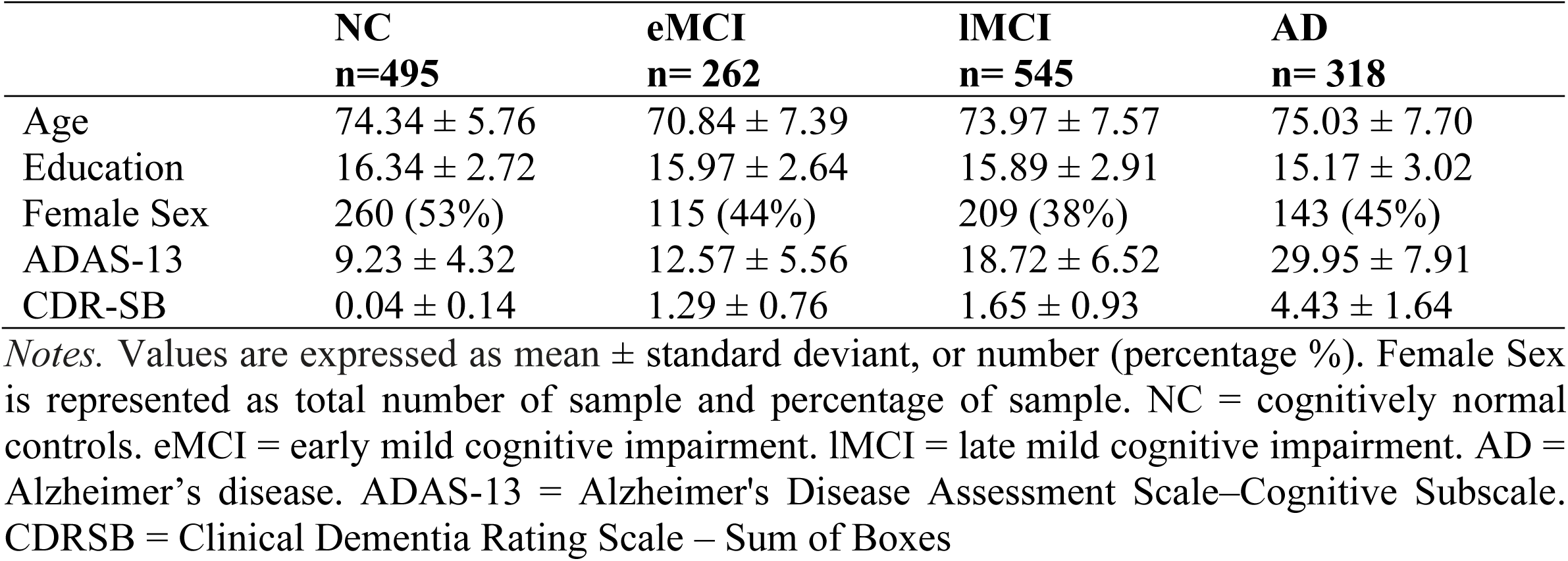
Demographic information for cognitively normal, early and late MCI, and AD participants.

|  | **NC**<br>**n=495** | **eMCI**<br>**n= 262** | **lMCI**<br>**n= 545** | **AD**<br>**n= 318** |
| --- | --- | --- | --- | --- |
| Age | 74.34 ± 5.76 | 70.84 ± 7.39 | 73.97 ± 7.57 | 75.03 ± 7.70 |
| Education | 16.34 ± 2.72 | 15.97 ± 2.64 | 15.89 ± 2.91 | 15.17 ± 3.02 |
| Female Sex | 260 (53%) | 115 (44%) | 209 (38%) | 143 (45%) |
| ADAS-13 | 9.23 ± 4.32 | 12.57 ± 5.56 | 18.72 ± 6.52 | 29.95 ± 7.91 |
| CDR-SB | 0.04 ± 0.14 | 1.29 ± 0.76 | 1.65 ± 0.93 | 4.43 ± 1.64 |

*Notes.* Values are expressed as mean ± standard deviant, or number (percentage %). Female Sex is represented as total number of sample and percentage of sample. NC = cognitively normal controls. eMCI = early mild cognitive impairment. lMCI = late mild cognitive impairment. AD = Alzheimer's disease. ADAS-13 = Alzheimer's Disease Assessment Scale–Cognitive Subscale. CDRSB = Clinical Dementia Rating Scale – Sum of Boxes

### 2.3 Structural MRI acquisition and processing

All participants were imaged using a 3T scanner with T1-weighted imaging parameters (see http://adni.loni.usc.edu/methods/mri-tool/mri-analysis/ for the detailed MRI acquisition protocol). Baseline scans were downloaded from the ADNI public website.

T1w scans for each participant were pre-processed through our standard pipeline including noise reduction ^20^, intensity inhomogeneity correction^21^ and intensity normalization into range [0-100]. The pre-processed images were then both linearly (9 parameters: 3 translation, 3 rotation, and 3 scaling) ^22^ and nonlinearly (1 mm^3^ grid) ^23^ registered to the MNI-ICBM152-2009c average ^24^. The quality of the linear and nonlinear registrations was visually verified by an experienced rater (author M.D.), blinded to diagnostic group. Only seven datasets did not pass this quality control step and were discarded.

### 2.4 SNIPE

Scoring by Nonlocal Image Patch Estimator (SNIPE) was used to measure the extent of AD-related change in the hippocampus using the linearly registered preprocessed T1-weighted images ^16,18^. SNIPE segmentations were visually verified by an experience rater (author N.S.) The SNIPE procedure used has been previously described in detail ^25^.

### 2.5 Data availability statement

The data used for this analysis are available on request from the ADNI database (ida.loni.usc.edu).

### 2.6 Statistical Analysis

Analyses were performed using ‘R’ software version 4.0.5. Linear regression models were conducted to determine whether hippocampal grading would influence cognitive scores (CDR-SB and ADAS-13). The model examined the association between *CognitiveScore* (ADAS-13 or CDR-SB) and Hippocampal *Grading* (Right and Left). Diagnosis was the categorical variable of interest, indicated by NC, eMCI, lMCI, or AD status with NC serving as the baseline. *Grading:Diagnosis* denotes an interaction term between Grading and Diagnosis, reflecting differences in the slope of Grading between the diagnostic groups. The models also included age, sex, and years of education as covariates, with the regression centered on SNIPE score = 0.0 as follows:

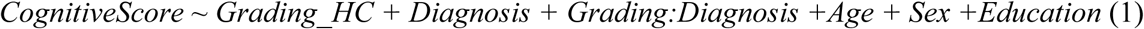

Correction of multiple comparisons was completed using false discovery rate (FDR); p-values are reported as raw values with significance determined by FDR correction marked in Table 2.

**Table 2:** Linear regression model results showing effects of grading on cognitive scores

|  | ADAS-13:Left HC | ADAS-13:Right HC | CDR-SB:Left HC | CDR-SB:Right HC |
| --- | --- | --- | --- | --- |
| Intercept | $\beta=15.69, t=7.57, p<.001^*$ | $\beta=18.17, t=8.73, p<.001^*$ | $\beta= 1.06, t= 3.12, p=.002^*$ | $\beta= 1.14, t=3.31, p<.001^*$ |
| Grading | $\beta=-6.16, t= -3.95, p<.001^*$ | $\beta= -7.04, t=-4.28, p<.001^*$ | $\beta= -0.21, t= -0.82, p=.41$ | $\beta= -0.21, t= -0.76, p=.45$ |
| Age | $\beta= -0.03, t= -1.11, p=.27$ | $\beta= -0.05, t=-2.02, p=.04$ | $\beta= -0.01, t= -2.49, p=.01^*$ | $\beta= -0.01, t= -2.53, p=.01^*$ |
| Male Sex | $\beta=0.88, t=2.98, p=.003^*$ | $\beta=0.87, t=3.00, p=.003^*$ | $\beta= -0.03, t= -0.71, p=.48$ | $\beta= -0.03, t= -0.63, p=.53$ |
| Education | $\beta= -0.21, t= -4.06, p<.001^*$ | $\beta= -0.24, t= -4.80, p<.001^*$ | $\beta= -0.02, t= -1.85, p=.06$ | $\beta= -0.02, t= -2.36, p=.02^*$ |
| eMCI | $\beta=4.19, t=5.83, p<.001^*$ | $\beta=4.10, t=5.35, p<.001^*$ | $\beta=1.28, t=10.96, p<.001^*$ | $\beta=1.30, t=10.25, p<.001^*$ |
| lMCI | $\beta=8.13, t=15.09, p<.001^*$ | $\beta=7.78, t=13.45, p<.001^*$ | $\beta=1.58, t=18.07, p<.001^*$ | $\beta=1.58, t=16.50, p<.001^*$ |
| AD | $\beta=17.61, t=28.80, p<.001^*$ | $\beta=16.65, t=25.49, p<.001^*$ | $\beta=4.08, t=40.96, p<.001^*$ | $\beta=4.06, t=37.48, p<.001^*$ |
| Grading*eMCI | $\beta= -5.06, t= -2.22, p=.027^*$ | $\beta= -4.54, t= -1.96, p=.05$ | $\beta= -0.32, t=-0.87, p=.38$ | $\beta= -0.36, t=-0.94, p=.35$ |
| Grading*lMCI | $\beta= -5.04, t= -2.74, p= .006^*$ | $\beta= -4.12, t= -2.23, p=.03^*$ | $\beta= -0.73, t= -2.43, p=.02^*$ | $\beta= -0.66, t= -2.22, p=.03^*$ |
| Grading*AD | $\beta= -3.94, t= -1.85, p= .06$ | $\beta= -5.31, t= -2.51, p=.01^*$ | $\beta= -1.84, t= -5.28, p<.001^*$ | $\beta= -1.58, t= -4.49, p<.001^*$ |
| Adjusted R-squared | 0.65 | 0.66 | 0.73 | 0.73 |

Note: Bolded values are statistically significant. \* Represents results that were significant after false discovery rate correction
for multiple comparisons. eMCI = early mild cognitive impairment; lMCI= late mild cognitive impairment; AD= Alzheimer's
Disease; HC = hippocampus; ADAS-13 = Alzheimer's Disease Assessment Scale-13; CDR-SB = Clinical Dementia Rating Scale –
Sum of Boxes.

## 3. Results

Table 1 presents the demographic information for each group. There was no significant difference in age between the NC and lMCI (*t*=0.9, *p*=0.4) or between NC and AD (*t*=1.36, *p*=0.2), but the eMCI group were 3.5y younger that NC (*t*=6.67, *p*<.001). There was no significant difference in education between NC and eMCI (*t*=1.83, *p*=.07), but NC had higher education than lMCI (*t*=2.58, *p*=.01) and AD (*t*=5.64, *p<*.001). As expected, the average ADAS-13 score increased from NC to AD. Statistically significant differences were observed between each successive stage of decline were observed, NC < eMCI < lMCI < AD (NC:eMCI, *t*=-8.45, *p*<.001; eMCI:lMCI, *t*=-13.89, *p*<.001; and lMCI:AD *t=*-21.43, *p*<.001). Similarly, CDR-SB increased with NC < eMCI < lMCI < AD with statistically significant differences between each successive group (NC:eMCI, *t*=-26.47, *p*<.001; eMCI:lMCI, *t*=-5.88, *p*<.001; and lMCI:AD *t=*-27.72, *p*<.001).

Table 2 summarizes the results of the linear regression models for both the ADAS-13 and CDR-SB analyses. Figure 1 shows a scatterplot of individual cognitive scores and hippocampal grading values for both the ADAS-13 and CDR-SB.

**Figure 1:**
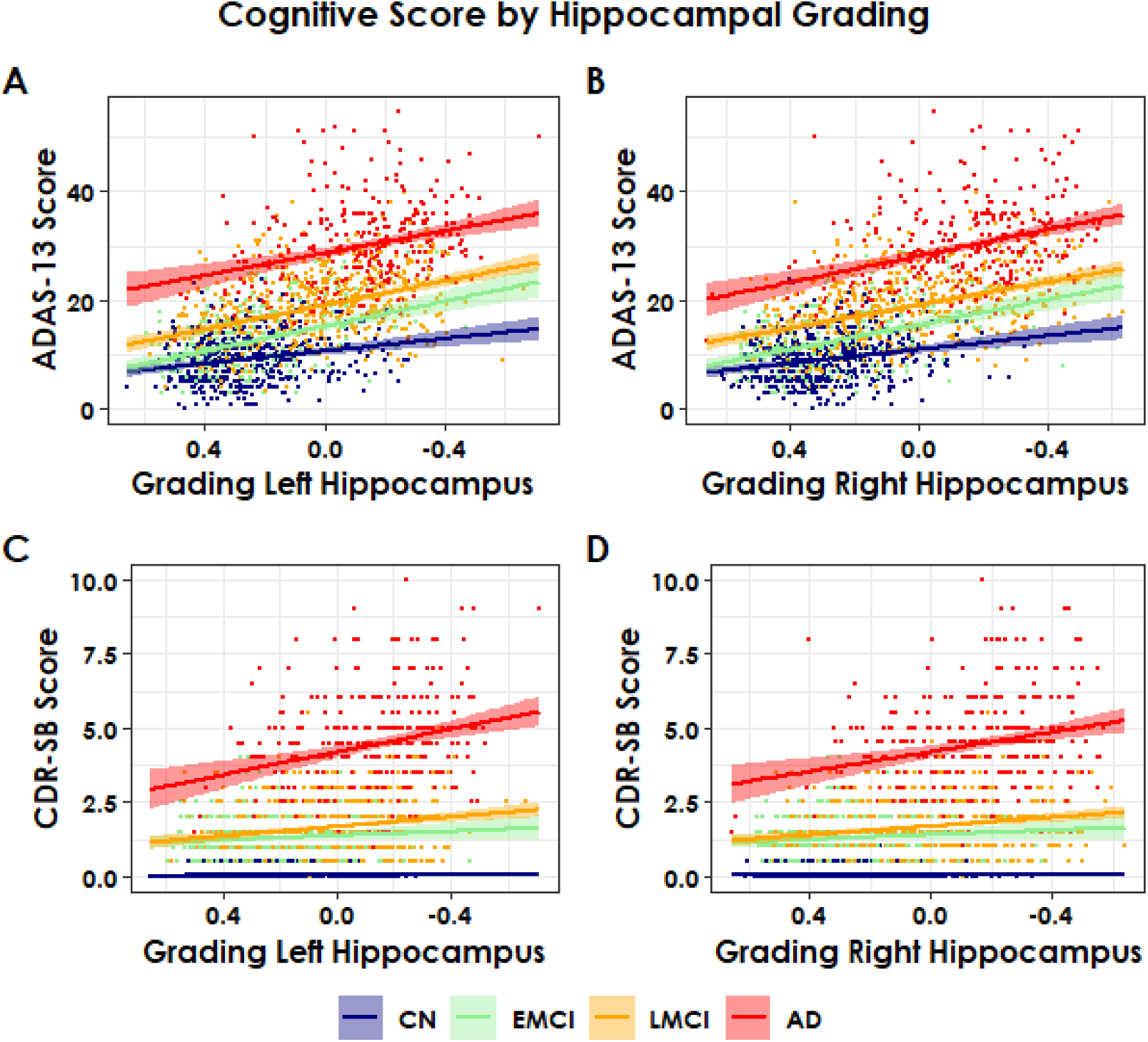
Scatterplots of Cognitive Score by Hippocampal Grading All images show individual points grouped by color as well as the regression line for each group. Dark blue lines = Cognitive normal (CN); Light green lines = early mild cognitive impairment (eMCI); orange lines = late mild cognitive impairment (lMCI); and red lines = Alzheimer’s disease (AD). ADAS-13 = Alzheimer’s Disease Assessment Scale–Cognitive Subscale. CDR-SB = Clinical Dementia Rating Scale – Sum of Boxes. Negative grading scores indicate greater similarity to the Alzheimer’s anatomy, while positive scores indicate similarity to healthy controls. From left to right, the x-axis starts with A&B) Higher ADAS-13 scores were associated with decreases in hippocampal grading in all groups. C&D) Higher CDR-SB scores were associated with decreases in hippocampal grading in all groups except CN.

### 3.1 ADAS-13 and Hippocampal Grading

For the left hippocampus (lHC) analysis, the overall effect of lHC grading on ADAS-13 scores in the NC group was significant (*t*= -3.95, *p*<.001), demonstrating that decreases in lHC grading were associated with increases in ADAS-13 scores. In addition, all patient groups had significantly greater intercepts for ADAS-13 than the NCs, and ADAS-13 scores progressively increased (i.e., lower performance) from NC to eMCI (4.19 points more than NC, *t*= 5.83, *p*<.001), lMCI (8.13 points more than NC, *t*= 15.09, *p*<.001), and AD (17.61 points more than NC, *t*= 28.80, *p*<.001) at the model center (where SNIPE grading = 0.0). Furthermore, the interaction between lHC grading and ADAS-13 was significant for eMCI (*t*= -2.22, *p*=.03) and lMCI (*t*= -2.74, *p*=.006), and marginally significant for AD (*t*= -1.85, *p*=.06); i.e., the slopes of changes in ADAS-13 scores associated with changes in lHC grading were significantly steeper in all disease cohorts in comparison with the NCs. The slope for eMCI and lMCI was almost twice that for NC. When examining the covariates, age was not associated with change in ADAS-13 scores (*t*= -1.11, *p*=.27). Male sex was associated with almost 1 point increase in ADAS-13 scores (*t*= 2.98, *p*=.003), whereas increased education was associated with slightly lower (−0.21 points) ADAS-13 scores (*t*= -4.06, *p*<.001). Overall, 65% of the variance in ADAS-13 scores (adjusted R^2^=0.65) can be explained by the variables included in this model.

For the right hippocampus (rHC) analysis, the overall effect of rHC grading on ADAS-13 scores in the NC group was significant (*t*= -4.28, *p*<.001), demonstrating that increases in ADAS-13 scores were associated with decreases in rHC grading. In addition, all patient groups had significantly greater intercepts for ADAS-13 than the NCs, and ADAS-13 scores progressively increased (i.e., lower performance) from NC to eMCI (4.10 points more than NC, *t*= 5.35, *p*<.001), lMCI (7.78 points more than NC, *t*= 13.45, *p*<.001), and AD (16.65 points more than NC, *t*= 25.49, *p*<.001) at the model center (where SNIPE grading = 0.0). Furthermore, the interaction between rHC grading and ADAS-13 was significant for eMCI (*t*= -1.96, *p*=.05), to lMCI (*t*= -2.23, *p*=.03), and to AD (*t*= -2.51, *p*=.01); i.e., the slopes of changes in ADAS-13 scores associated with changes in rHC grading were significantly steeper in all disease cohorts in comparison with the NCs. When examining the covariates, increased age was associated with slight increases in ADAS-13 scores (0.05 points, *t*= 2.02, *p*=.04). Male sex was associated with almost 1 point increase in ADAS-13 scores (*t*= 3.00, *p*=.003), whereas increased education was associated with slightly lower ADAS-13 scores (−0.24 points, *t*= -4.80, *p*<.001). Overall, 66% of the variance in ADAS-13 scores (adjusted R^2^=0.66) can be explained by the variables included in this model.

### 3.2 CDR-SB and Hippocampal Grading

For the left hippocampus model, the effect of lHC grading on CDR-SB scores was not significant (*t=* -0.82, *p*=.41) for the NC group. All patient groups had significantly greater intercepts for CDR-SB than the NC group at model center, and CDR-SB scores progressively increased (i.e., lower performance) from NC to eMCI (1.28 points more than NC, *t*= 10.96, *p*<.001), to lMCI (1.58 points more than NC, *t*= 18.07, *p*<.001), and to AD (4.08 points more than NC, *t*= 40.90, *p*<.001). The interaction between lHC grading and CDR-SB was not significant for eMCI (*t*= -0.87, *p*=.38), but was significant for lMCI (*t*= -2.43, *p*=.02) and AD (*t*= -5.28, *p*<.001); i.e., the slopes of changes in CDR-SB scores associated with changes in HC grading were significantly steeper slopes in the later disease cohorts (lMCI & AD) compared to NCs. When examining the covariates, increased age was associated with increases in CDR-SB scores (0.01 points, *t*= 2.49, *p*=.01), male sex was not associated with change in CDR-SB scores (*t*= - 0.71, *p*=.48), and education was only marginally associated with lower CDR-SB scores (−0.02 points, *t*= -1.85, *p*=.06). Overall, 73% of the variance in CDR-SB scores (adjusted R^2^=0.73) can be explained by the variables included in this model.

For the right hippocampus model, the effect of rHC grading on CDR-SB scores was not significant (*t*= -0.76, *p*=.45) for the NC group. All patient groups had significantly greater intercepts for CDR-SB than the NCs, and CDR-SB scores progressively increased (i.e., lower performance) from NC to eMCI (1.30 points over NC, *t*= 10.25, *p*<.001), to lMCI (1.58 points over NC *t*= 16.50, *p*<.001, and to AD (4.06 points more than NC, *t*= 37.48, *p*<.001). The interaction between rHC grading and CDR-SB was not significant for eMCI (*t*= -0.94, *p*=.35), but was significant for lMCI (*t*= -2.22 *p*=.03), and AD (*t*= -4.49, *p*<.001); i.e., the slopes of changes in CDR-SB scores associated with changes in HC grading were significantly different (steeper slopes) in the later disease cohorts (lMCI & AD) in contrast with the NCs. When examining the covariates, increased age was associated with slight increases in CDR-SB scores (0.01 points, *t*= 2.53, *p*=.01), male sex was not associated with change in CDR-SB scores (*t*= - 0.63, *p*=.53), whereas increased education was associated with slightly lower CDR-SB scores (0.02 points, *t*= -2.36, *p*=.02). Overall, 73% of the variance in CDR-SB scores (adjusted R^2^=0.73) can be explained by the variables included in this model.

## 4. Discussion

The current study used SNIPE to measure hippocampal grading in NC, eMCI, lMCI, and AD, and compared these grading scores to global cognitive functioning as measured by the ADAS-13 and CDR-SB. In contrast to NCs, both lMCI and AD required less hippocampal change to have decreased ADAS-13 and CDR-SB scores (reflected by their significantly steeper slopes in the regression models). The eMCI group required less hippocampal change compared to normal controls to have decreases in only the ADAS-13; whereas this group’s association between hippocampal change and CDR-SB scores did not differ from NC. These findings indicate a relationship between lower cognitive scores in eMCI, lMCI, and AD with decreases in hippocampal grading.

Previous associations between global cognition in people with MCI and hippocampal volume have been mixed. Vipin et al. (2018) reported no relationship between hippocampal volume and global cognition in MCI^14^, while Peng et al. (2014) found an association between hippocampal volume and global cognition in people with MCI^15^. These conflicting findings could be associated with the sensitivity of the MMSE to early cognitive decline as well as the use of hippocampal volume.

In the current study, we observed an association between hippocampal grading and global cognition, as measured by the ADAS-13 score in all groups - NC, eMCI, lMCI, and AD. This finding suggests that SNIPE hippocampal grading is sensitive to global cognitive declines due to aging (in the NC group), early in the disease process (i.e., eMCI), and to the progressive changes that occur later in AD-related pathology. Taken together with the previous research on SNIPE ^16–18^, these results suggest that this method could be useful in the future prediction of cognitive decline and diagnostic status early in the disease trajectory.

When examining global cognition using the CDR-SB, associations between hippocampal grading were only observed in the lMCI and AD groups. As expected, CDR-SB scores were low for the normal controls (i.e., either 0 or 0.5), resulting in a flat slope for this group. As can be seen in Figure 1, the eMCI group has a much smaller range and median (0-4; median=1) CSR-SB values than both lMCI (0-5.5; median=1.5) and AD (1-10; median=5.5). The lack of association with the eMCI group may thus be related to the limited range of CDR-SB scores in people with eMCI. The relationship became stronger with lMCI and AD, showing that the relationship between hippocampal grading and CDR-SB is stronger in those with more severe declines. Consistent with previous studies, associations between CDR-SB and hippocampal grading were also stronger in the left, rather than right hippocampus ^15,26^.

While both ADAS-13 and CDR-SB were associated with hippocampal grading for both lMCI and AD, only the ADAS-13 was associated with hippocampal grading in eMCI and NC. CDR-SB was more sensitive (than ADAS-13) to changes in hippocampal grading that occur later in the disease progression, as reflected by the interaction with grading and lMCI and AD. These findings suggest that the relationship between grading and cognitive score can be sensitive to different stages of disease progression.

Regarding the normal controls, these findings suggest they would require much more hippocampal change (as measured by grading) for the same amount of cognitive decline to occur compared to the patient groups. For example, HC changes may affect the grading score, but compensation through cognitive reserve or plasticity may limit declines in cognitive functioning measured by the cognitive tests. Further research is needed to elucidate this issue.

The present study has a few limitations that should be investigated in future research. All participants in our sample had a high education, which may limit the interpretation and generalizability of the results to other populations. To improve our sample size, participants were included from ADNI-1, ADNI -2, and ADNI -GO. Therefore, we could not differentiate between cognitively normal participants with and without subjective cognitive decline (SCD) because a question to measure this construct was only introduced for ADNI-2. Recent research has observed that hippocampal volume decreases are associated with episodic memory performance in people with SCD ^27^. Thus, future research should determine whether SNIPE grading is sensitive to hippocampal changes and global cognitive functioning in people with SCD. These findings should be replicated in longitudinal data to determine whether grading scores are predictive of clinical cognitive change at an individual level.

## Conclusion

The findings from this study indicate a strong association between hippocampal grading changes and global cognition. Importantly, the relationships were observed not only late in the disease course (AD and lMCI) but also earlier in the course of the disease during eMCI. Hippocampal grading may be a useful measure that is sensitive to progressive changes in global cognition as measured by the ADAS-13 starting in the preclinical phase and CDR-SB starting in the prodromal phase of AD. Future work is needed to determine if HC grading is predictive and associated with longitudinal changes in cognition at an individual level.

## Data Availability

A written request was submitted to ADNI and after approval access was granted data was downloaded from adni. Therefore all data used in this manuscript can be downloaded from adni upon request.

## Acknowledgments

Data collection and sharing for this project was funded by the Alzheimer’s Disease Neuroimaging Initiative (ADNI) (National Institutes of Health Grant U01 AG024904) and DOD ADNI (Department of Defense award number W81XWH-12-2-0012). ADNI is funded by the National Institute on Aging, the National Institute of Biomedical Imaging and Bioengineering, and through generous contributions from the following: AbbVie, Alzheimer’s Association; Alzheimer’s Drug Discovery Foundation; Araclon Biotech; BioClinica, Inc.; Biogen; Bristol-Myers Squibb Company; CereSpir, Inc.; Cogstate; Eisai Inc.; Elan Pharmaceuticals, Inc.; Eli Lilly and Company; EuroImmun; F. Hoffmann-La Roche Ltd and its affiliated company Genentech, Inc.; Fujirebio; GE Healthcare; IXICO Ltd.; Janssen Alzheimer Immunotherapy Research & Development, LLC.; Johnson & Johnson Pharmaceutical Research & Development LLC.; Lumosity; Lundbeck; Merck & Co., Inc.; Meso Scale Diagnostics, LLC.; NeuroRx Research; Neurotrack Technologies; Novartis Pharmaceuticals Corporation; Pfizer Inc.; Piramal Imaging; Servier; Takeda Pharmaceutical Company; and Transition Therapeutics. The Canadian Institutes of Health Research is providing funds to support ADNI clinical sites in Canada. Private sector contributions are facilitated by the Foundation for the National Institutes of Health (www.fnih.org). The grantee organization is the Northern California Institute for Research and Education, and the study is coordinated by the Alzheimer’s Therapeutic Research Institute at the University of Southern California. ADNI data are disseminated by the Laboratory for Neuro Imaging at the University of Southern California.

## References

1. Petersen RC. Mild cognitive impairment as a clinical entity and treatment target. Arch Neurol. 2005;62(7):1160–1163; discussion 1167. doi:10.1001/archneur.62.7.1160

2. Association Association. What is Alzheimer’s disease. https://www.alz.org/alzheimers-dementia/what-is-alzheimers. Published 2021.

3. Bettio LEB, Rajendran L, Gil-Mohapel J. The effects of aging in the hippocampus and cognitive decline. Neurosci Biobehav Rev. 2017;79:66–86. doi:10.1016/j.neubiorev.2017.04.030

4. Jack Jr CR, Bennett DA, Blennow K, Carrillo MC, Dunn B, Haeberlein SB, Holtzman DM, Jagust W, Jessen F, Karlawish J, Liu E.. NIA-AA Research Framework: Toward a biological definition of Alzheimer’s disease. Physiol Behav. 2018;176(5):139–148. doi:10.1016/j.jalz.2018.02.018.NIA-AA

5. Fjell AM, McEvoy L, Holland D, Dale AM, Walhovd KB. What is normal in normal aging? Effects of aging, amyloid and Alzheimer’s disease on the cerebral cortex and the hippocampus. Prog Neurobiol. 2014;117:20–40. doi:10.1016/j.pneurobio.2014.02.004

6. Jack CR, Wiste HJ, Weigand SD, et al. Age, sex and APOE ε4 effects on memory, brain structure and βamyloid across the adult lifespan. JAMA Neurol. 2015;72(5):511–519. doi:10.1001/jamaneurol.2014.4821.Age

7. Reisberg B, Shulman MB, Torossian C, Leng L, Zhu W. Outcome over seven years of healthy adults with and without subjective cognitive impairment. 2010;6:11–24. doi:10.1016/j.jalz.2009.10.002

8. Köhncke Y, Düzel S, Sander MC, Lindenberger U, Kühn S, Brandmaier AM. Hippocampal and Parahippocampal Gray Matter Structural Integrity Assessed by Multimodal Imaging Is Associated with Episodic Memory in Old Age. 2021;31(3):1464–1477. doi:10.1093/cercor/bhaa287

9. Persson J, Pudas S, Lind J, Kauppi K. Longitudinal Structure--Function Correlates in Elderly Reveal MTL Dysfunction with Cognitive Decline. 2012;22(10):2297–2304. doi:10.1093/cercor/bhr306

10. Gorbach T, Pudas S, Lundquist A, et al. Longitudinal association between hippocampus atrophy and episodic-memory decline. Neurobiol Aging. 2017;51:167–176. doi:10.1016/j.neurobiolaging.2016.12.002

11. Schneider ALC, Senjem ML, Wu A, et al. Neural correlates of domain-specific cognitive decline. Neurology. 2019; 92(10),e1051–e1063 doi:10.1212/WNL.0000000000007042

12. Yang C, Ren J, Li W, Lu M, Wu S, Chu T. Brain and Cognition Individual-level morphological hippocampal networks in patients with Alzheimer ‘s disease.2021;151:105748.

13. Dawe RJ, Bennett DA, Yu L, Arfanakis K, Boyle PA, Schneider JA. Late-life cognitive decline is associated with hippocampal volume, above and beyond its associations with traditional neuropathologic indices. 2020;61(1):209–218. doi:10.1002/alz.12009

14. Vipin A, Jing H, Foo L, Kai J, Lim W, Jude R. Regional White Matter Hyperintensity Influences Grey Matter Atrophy in Mild Cognitive Impairment. 2018;66(2):533–549. doi:10.3233/JAD-180280

15. Peng G, Feng Z, He F, et al. Correlation of Hippocampal Volume and Cognitive Performances in Patients with Either Mild Cognitive Impairment or Alzheimer ‘ s disease. 2015;21(1):15–22. doi:10.1111/cns.12317

16. Coupé P, Eskildsen SF, Manjón JV., et al. Scoring by Nonlocal Image Patch Estimator for early detection of Alzheimer’s disease. NeuroImage Clin. 2012;1(1):141–152. doi:10.1016/j.nicl.2012.10.002

17. Coupe P, Fonov VS, Bernard C, et al. Detection of Alzheimer ‘ s Disease Signature in MR Images Seven Years Before Conversion to Dementia: Toward an Early Individual Prognosis. 2015;36(12):4758–4770. doi:10.1002/hbm.22926

18. Coupé P, Eskildsen SF, Manjón JV, Fonov VS, Collins DL. Simultaneous segmentation and grading of anatomical structures for patient ‘ s classi fi cation : Application to Alzheimer’ s disease. Neuroimage. 2012;59(4):3736–3747. doi:10.1016/j.neuroimage.2011.10.080

19. Mitchell AJ. A meta-analysis of the accuracy of the mini-mental state examination in the detection of dementia and mild cognitive impairment. J Psychiatr Res. 2009;43(4):411–431. doi:10.1016/j.jpsychires.2008.04.014

20. Coupe P, Yger P, Prima S, Hellier P, Kervrann C, Barillot C. An optimized blockwise nonlocal means denoising filter for 3-D magnetic resonance images. IEEE Trans Med Imaging. 2008;27(4):425–441. doi:10.1109/TMI.2007.906087

21. Sled JG, Zijdenbos AP, Evans AC. A nonparametric method for automatic correction of intensity nonuniformity in mri data. IEEE Trans Med Imaging. 1998;17(1):87–97. doi:10.1109/42.668698

22. Dadar M, Fonov VS, Collins DL. A comparison of publicly available linear MRI stereotaxic registration techniques. Neuroimage. 2018;174:191–200. doi:10.1016/j.neuroimage.2018.03.025

23. Avants BB, Epstein CL, Grossman M, Gee JC. Symmetric diffeomorphic image registration with cross-correlation: Evaluating automated labeling of elderly and neurodegenerative brain. Med Image Anal. 2008;12(1):26–41. doi:10.1016/j.media.2007.06.004

24. Fonov V, Evans AC, Botteron K, Almli CR, McKinstry RC, Collins DL. Unbiased average age-appropriate atlases for pediatric studies. Neuroimage. 2011;54(1):313–327. doi:10.1016/j.neuroimage.2010.07.033

25. Dadar M, Gee M, Shuaib A, Duchesne S, Camicioli R. Cognitive and motor correlates of grey and white matter pathology in Parkinson’s disease. NeuroImage Clin. 2020;27:102353. doi:10.1016/j.nicl.2020.102353

26. Basso M, Yang J, Warren L, et al. Volumetry of amygdala and hippocampus and memory performance in Alzheimer’s disease. Psychiatry Res - Neuroimaging. 2006;146(3):251–261. doi:10.1016/j.pscychresns.2006.01.007

27. Caillaud M, Hudon C, Boller B, et al. Evidence of a relation between hippocampal volume, white matter hyperintensities, and cognition in subjective cognitive decline and mild cognitive impairment. Journals Gerontol-Ser B Psychol Sci Soc Sci. 2019;75(7):1382–1392. doi:10.1093/geronb/gbz120

